# Impact of School-Led Total Sanitation on health outcomes among children aged 6-59 months in Baringo County, Kenya: A quasi-experimental study

**DOI:** 10.64898/2026.07.14.26358036

**Authors:** Phanice Kerubo Omari, Zachary Masimba Ondicho, Simon Muturi Karanja, Susan Njoki Mambo

**Author notes:** Corresponding author: Phanice Kerubo Omari.

## Abstract

Diarrheal disease remains a leading cause of morbidity and mortality among children under five globally, with poor sanitation and hygiene accounting for over 88% of diarrhea and malnutrition burden. In Kenya, diarrhea ranks third in under-five mortality, particularly affecting arid and semi-arid regions. School-Led Total Sanitation (SLTS), adapted from Community-Led Total Sanitation (CLTS), uses pupils as change agents for household hygiene knowledge transfer. However, SLTS effectiveness in addressing diarrhea and malnutrition has not been evaluated in Kenya. This study assessed SLTS effects on diarrheal disease and nutritional outcomes among children aged 5-59 months in Baringo County. A pre- and post-test quasi-experimental design with nonequivalent control groups was employed in Mogotio (intervention) and Baringo South (control) sub-counties. Using multistage sampling, 440 children aged 6-59 months were enrolled. The six-month SLTS intervention included capacity building, school health club formation, triggering activities using Participatory Rural Appraisal tools, Information, Education, and Communication materials distribution, continuous sensitization, and household monitoring. Data were collected at baseline and three months post-intervention using electronic questionnaires and anthropometric measurements. Nutritional status was assessed using WHO Anthro software z-scores for length/height-for-weight (HWZ), and weight-for-age (WAZ) to determine wasting, and underweight prevalence. Chi-square analysis assessed intervention-control differences. Baseline and endline socio-demographic characteristics were comparable between groups. At endline, no significant nutritional outcome differences were observed: wasting prevalence was at 15.0% versus 16.4% (χ²=0.155, df=1, p=0.694) while underweight was 12.3% versus 13.6% (χ²=0.181, df=1, p=0.670). However, diarrheal disease prevalence significantly reduced in intervention versus control groups: 5.9% versus 13.2% (χ²=6.738, df=1, p=0.009), representing a 53% reduction. SLTS intervention showed no significant effect on nutritional outcomes but demonstrated a significant reduction in diarrheal disease among children aged 6-59 months. These findings provide strong evidence for integrating school-based sanitation and hygiene approaches into broader public health strategies addressing diarrheal diseases.

## 1.0 Introduction

Diarrheal disease remains a major cause of morbidity and mortality among children under five years globally, accounting for an estimated 444,000 deaths annually, with 90% of these occurring in Asia and Africa[1,2] In Kenya, diarrhea is the third leading cause of under-five mortality, contributing to more than 4,600 deaths each year [3]. The burden is not evenly distributed; children living in arid and semi-arid regions and informal settlements face a disproportionately high risk of diarrheal morbidity and mortality [4]. In Baringo County, diarrhea ranks fourth among the top ten causes of morbidity for both under-fives and the general population [5].

Poor sanitation and hygiene account for more than 60% of the disease burden from diarrhea and malnutrition [6]. The relationship between diarrhea and malnutrition is bidirectional: recurrent diarrheal episodes predispose children to malnutrition, while malnourished children are more susceptible to infectious diseases due to compromised immunity[7,8]. Globally, 23.2% of children under five are stunted and 6.6% are wasted, with sub-Saharan Africa and South Asia accounting for more than half of these cases. In Kenya, the prevalence of stunting, wasting, and underweight among children under five is 18%, 5%, and 10%, respectively, while in Baringo County, these rates are even higher—21%, 14%, and 20%—underscoring the urgent need for targeted interventions [9].

Sanitation and hygiene interventions have consistently been shown to reduce diarrhea incidence. Meta-analyses and intervention trials indicate that sanitation and hygiene interventions can reduce diarrheal prevalence by 23–69% [10,11]. However, their impact on nutritional outcomes remains mixed. While some studies have reported improvements in child growth indicators following such interventions [12], others have found minimal or no effect [13–15].

School-Led Total Sanitation (SLTS), adapted from the Community-Led Total Sanitation (CLTS) approach, is one intervention designed to improve sanitation and hygiene behaviors among pupils and their communities [16]. By engaging pupils as change agents, SLTS promotes the transfer of hygiene knowledge and practices to households, potentially benefiting younger siblings and other community members. Despite its potential, the effectiveness of SLTS in addressing both diarrhea and malnutrition has not been evaluated in Kenya. This study therefore assessed the effect of SLTS on diarrheal disease and nutritional outcomes among children under five years in Baringo County.

## 2.0 Materials and methods

### 2.1 Study setting

This study was conducted in Baringo County, Kenya, one of the 47 devolved administrative units in the country, located in the Rift Valley region. The county covers approximately 11,015 km² and has an estimated population of 754,014 people [17]. It is largely characterized by arid and semi-arid climatic conditions. Two sub-counties, Mogotio (intervention site) and Baringo South (control site), were purposively selected based on comparable socio-demographic characteristics and high diarrhea and malnutrition prevalence.

### 2.2 Study population

The study sample consisted of caregivers with at least one child aged 6-59 months and with at least one child from the participating schools

### 2.3 Sample size calculation

This study is part of a major study that aimed to evaluate the effect of school-led total sanitation on pupils, caregivers’ hygiene and sanitation knowledge and practices, and also on under-five health outcomes. The Fleiss method sample size formula[18] was used to determine the sample size for the study.

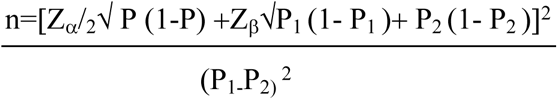

The sample size per study arm was 220, making a total of 440 participants.

### 2.4 Study design and sampling procedure

A pre-and post-test quasi-experimental with a nonequivalent control group design was adopted for this study. The study had an intervention (Mogotio sub-county) and control arm (Baringo South sub-county). The control site was selected based on the following criteria: - It is located in Baringo county with similar geographical, climatic, socio-economic and demographic characteristics as the intervention site, no active (Water, Sanitation and Hygiene) WASH interventions were going on or planned in the subcounty during the study period and the distance between the 2 sites is about 75 kilometers, which is a sufficient distance to minimize spillover effects or contamination.

Using a multistage cluster sampling method, three primary schools were selected from each of the two sub-counties. In the second stage, households were identified within the catchment areas of these schools. Caregivers in the households were eligible if they had at least one child aged 6–59 months and at least one school-aged child enrolled in one of the selected schools. In total, 440 respondents were selected across both the intervention and control arms. Eligible respondents were recruited through home visits. The baseline study was conducted between 19^th^ January and 27^th^ February 2022; the intervention lasted for six months from April 2022 to September 2022. There was a 3-month post-intervention stay period, and the endline assessment was conducted between 15^th^ January and 24^th^ February 2023.

### 2.5 Description of the SLTS intervention

The School-Led Total Sanitation (SLTS) intervention included the following components:

- **Capacity building:** A multidisciplinary team consisting of teachers, School Board of Management members, Parents Teachers’ Association representatives, Community Health Volunteers, and the area Public Health Officer was trained on SLTS objectives, implementation strategies, and monitoring procedures.
- **Formation of school health clubs:** Clubs were established in each participating school, comprising pupils from Grades 3–8. Members received training on topics including faecal contamination pathways, critical times for handwashing, safe faeces disposal, menstrual hygiene, and environmental sanitation.
- **Triggering activities:** Participatory Rural Appraisal (PRA) tools adapted from the CLTS approach[16] were used. Activities included school mapping, transect walks, F-diagram analysis, and faeces quantification exercises.
- **Information, Education, and Communication (IEC) Materials:** Branded T-shirts for club members, household brochures, thematic posters, and hygiene promotion messages painted on school latrine walls were distributed to reinforce key sanitation messages.
- **Continuous sensitization:** Hygiene messages were integrated into classroom lessons and reinforced during assemblies, especially before school breaks.
- **Household monitoring:** Weekly home visits were conducted by the school health team, Community Health Volunteers, and the principal investigator to monitor SLTS uptake and provide feedback to the school health clubs.

### 2.6 Data collection tools and procedures

Quantitative data were collected using an electronic questionnaire programmed in KoBo Collect. Trained interviewers administered the questionnaire to caregivers to obtain information on the incidence of diarrhoea in children under five during the two weeks preceding the survey. Diarrhea was defined as the passage of three or more watery stools in a day during the diarrhea assessment period, which was 2 weeks before the study.

Anthropometric assessments for children aged 6–59 months followed WHO standard procedures [19].

- **Height/Length:** Recumbent length was measured for children aged 6–23 months and for older children unable to stand, using a portable length board. Standing height for children aged 24–59 months was measured using the same type of board. Children were positioned according to WHO guidelines, with measurements recorded to the nearest 0.1 cm.
- **Weight:** Measured with a calibrated digital SECA® electronic weighing scale placed on a flat surface and zeroed before each use. Children were weighed barefoot, wearing light clothing, and measurements were recorded to the nearest 0.1 kg.
- **Mid-Upper Arm Circumference (MUAC):** Measured on the left arm using a non-stretch, colour-coded MUAC tape, placed midway between the acromion and olecranon processes. Measurements were recorded to the nearest 0.1 cm.

All enumerators underwent intensive training and standardization exercises. Measurement protocols were supervised daily, and equipment was calibrated regularly. Duplicate measurements were taken, and a third measurement was obtained if discrepancies exceeded 0.2 cm for height/length or MUAC, or 0.2 kg for weight.

### 2.7 Data analysis

Quantitative data were analyzed using IBM SPSS Statistics version 25. Normality of continuous variables was assessed using the Kolmogorov–Smirnov test. Missing data were handled using listwise deletion. Continuous variables were summarized as means and standard deviations, while categorical variables were expressed as frequencies and percentages. Group differences in categorical variables were evaluated using Pearson’s Chi-square test, with statistical significance set at p≤0.05.

Anthropometric indices were calculated using WHO Anthro software version 3.2.2. Weight-for-height z-scores (WHZ), and weight-for-age z-scores (WAZ) were computed to assess wasting and underweight, respectively, according to the WHO Child Growth Standards [20].

- **Wasting:** WHZ < –2 SD
- **Underweight:** WAZ < –2 SD

Children with z-scores ≥ –2 SD were classified as having normal nutritional status for that indicator. Implausible values outside WHO biologically plausible ranges (HAZ < –6 or > +6; WHZ < –5 or > +5; WAZ < –6 or > +5) were excluded.

### 2.8 Ethical considerations

Approval to conduct the study was obtained from the Jomo Kenyatta University of Agriculture and Technology Board of Postgraduate Studies (BPS-JKUAT), the University of Eastern Africa Baraton Ethics Review Board (ERB-UEAB), and the National Commission for Science, Technology, and Innovation (NACOSTI). Additional permissions were obtained from the Baringo County Government, County Department of Education, and County Department of Health. Written informed consent was obtained from caregivers. Confidentiality and anonymity were maintained throughout the study.

### 3.0 Results

### 3.1. Socio-demographic characteristics of the respondents

Socio-demographic characteristics of the respondents are shown in the table 1 below

**Table 1:**
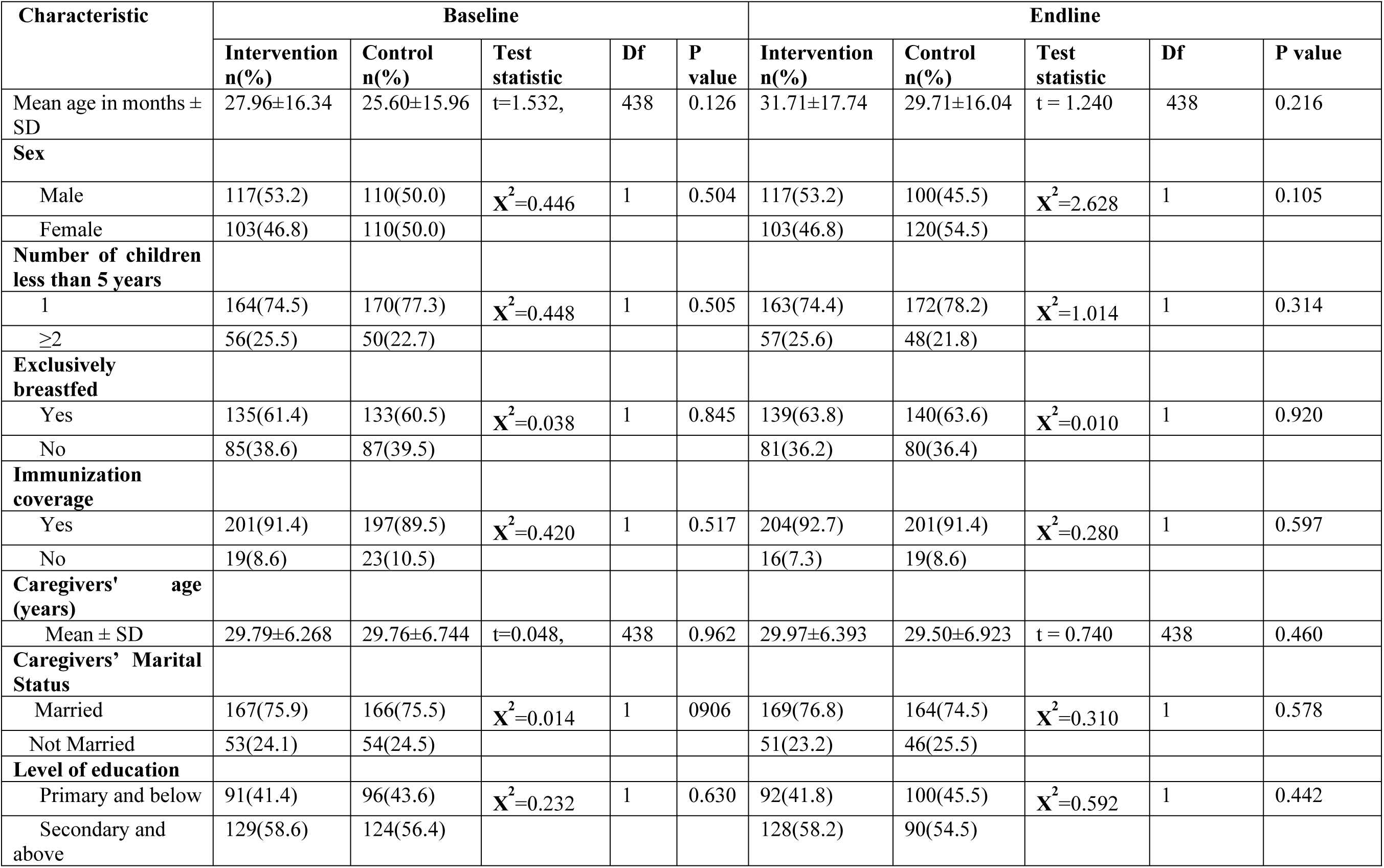
Socio-demographic characteristics of the respondents.

A total of 440 under-five children were enrolled at baseline, with 220 participants in both the intervention and control arms. At baseline, the mean age of children in the intervention group was 27.96 ± 16.34 months, compared to 25.60±15.96 months in the control group. At endline, the mean ages were slightly higher in both groups: 31.71±17.74 months in the intervention arm and 29.71 ± 16.04 months in the control arm.

The sex distribution was balanced across study arms, with males comprising 53.2% in the intervention group and 50.0% in the control group at baseline. At endline, sex distribution remained similar, with the intervention group having 53.2% (117) male and 46.8% (103) female children, and the control group having 45.5% (100) male and 54.5% (120) female children.

Other characteristics, such as exclusive breastfeeding, immunization coverage, caregivers’ mean age, marital status, and level of education, were comparable between the two groups at both baseline and endline. At baseline, 61.4% (135) of children in the intervention group were exclusively breastfed, compared to 60.5% (133) in the control group. The mean age of caregivers was also similar, with the intervention group at 29.79 ± 6.268 years and the control group at 29.76 ± 6.744 years. The majority of caregivers were married at baseline, accounting for 75.9% (167) in the intervention arm and 75.9% (166) in the control arm (Table 1).

### 3.2. Nutritional and health outcomes

Nutritional and health outcomes at baseline and endline evaluations were as presented in Table 2 below

**Table 2:** Comparison of child health outcomes of children aged 6-59 months Baseline and at endline following SLTS intervention in Baringo County Kenya.

| Indicator | Baseline |  |  |  |  | Endline |  |  |  |  |
| --- | --- | --- | --- | --- | --- | --- | --- | --- | --- | --- |
|  | Intervention<br>n(%) | Control<br>n(%) | X<sup>2</sup> | df | p-Value | Intervention<br>n(%) | Control<br>n(%) | X<sup>2</sup> | df | p-Value |
| **Weight -for-height Z-scores** |  |  |  |  |  |  |  |  |  |  |
| <-2.00 Wasted | 37(16.8) | 41(18.6) | 0.250 | 1 | 0.617 | 33(15.0) | 36(16.4) | 0.155 | 1 | 0.694 |
| ≥ 2.0: Normal | 183(83.2) | 179(81.4) |  |  |  | 187(85.0) | 184(83.6) |  |  |  |
| **Weight-for-age Z-score** |  |  |  |  |  |  |  |  |  |  |
| <-2.0:<br>Underweight | 36(16.4) | 31(14.1) | 0.442 | 1 | 0.506 | 27(12.3) | 30(13.6) | 0.181 | 1 | 0.670 |
| ≥ 2.0: Normal | 184(83.6) | 189(89.5) |  |  |  | 193(87.7) | 190(86.4) |  |  |  |
| **Diarrhoea in the last 2 weeks** |  |  |  |  |  |  |  |  |  |  |
| Yes | 28 (12.7) | 30(13.6) | 0.079 | 1 | 0.779 | 13(5.9) | 29(13.2) | 6.738 | 1 | 0.009 |
| No | 192(87.3) | 190(86.4) |  |  |  | 207(94.1) | 191(86.8) |  |  |  |

At baseline, the prevalence of wasting (WHZ < –2 SD) was 16.8% in the intervention arm and 18.6% in the control arm. At endline, wasting prevalence was 15.0% and 16.4% (WHZ < –2 SD), respectively. There was a slight decrease in the intervention group from 16.8% to 15.0%, while the control group showed a marginal reduction from 18.6% to 16.4%. Underweight prevalence (WAZ < –2 SD) dropped from 16.4% to 12.3% in the intervention arm and remained relatively unchanged in the control group (14.1% to 13.6%). There were no statistically significant differences between the intervention and control arms in wasting or underweight prevalence at either baseline or endline (p >0.05 for all comparisons).

A statistically significant reduction in diarrhea prevalence was observed in the intervention arm at endline compared to the control arm (5.9% vs. 13.2%, *X*^2^=6.738, df=1, p=0.009). No significant differences in diarrhea prevalence were observed at baseline (*X*^2^=0.0079, df=1p=0.779) (Table 2)

## 4.0 Discussion

This study found that the socio-demographic characteristics of the intervention and control groups were comparable at baseline, confirming the appropriateness of the selected sites for comparison. The lack of significant differences in factors such as age, sex, caregiver education, and marital status at the start of the study suggests that any observed differences in health outcomes at endline are likely attributable to the intervention rather than pre-existing disparities.

The prevalence of wasting showed minimal changes across both study arms. In the intervention group, wasting decreased slightly from 16.8% to 15.0%, while in the control group, it decreased from 18.6% to 16.4%. These differences were not statistically significant at either baseline (*X*^2^=0.250, df=1, p=0.617) or endline (*X*^2^=0.155, df=1. p=0.694).

A comprehensive analysis published in *Nature* demonstrated that wasting is a highly dynamic process of onset and recovery, with incidence patterns that may not be easily modified by single-component interventions[21]. The Chad study examining WASH (Water Sanitation and Hygiene) interventions for severe acute malnutrition similarly found that the interventions have measurable benefits on stunting but not on wasting[22]. A systematic review with meta-analysis found mixed evidence on the effects of WASH interventions on wasting indicators, particularly in community-based studies with short follow-up periods [23].

The limited impact on wasting may reflect the acute nature of this condition, which is primarily driven by recent illness episodes or inadequate food intake rather than long-term environmental improvements. The six-month intervention period may have been insufficient to establish sustained behavior change and environmental improvements necessary to prevent the infectious diseases that contribute to acute malnutrition [24]. Additionally, wasting is heavily influenced by seasonal patterns of food availability and disease transmission [25], factors that may not be adequately addressed by a single sanitation and hygiene intervention.

The intervention group showed improvement in underweight prevalence, decreasing from 16.4% to 12.3%, while the control group showed minimal change from 14.1% to 13.6%. However, these differences were not statistically significant (baseline *X*^2^=0.442, df=1, p=0.506; endline *X*^2^=0.181, df=1, p=0.670). Similar patterns have been observed in other school-based sanitation interventions. A systematic review examining single and combined WASH interventions found modest improvements in weight-for-age indicators, though statistical significance was not achieved in all study sites[23]. The implications from the major WASH trials suggest that weight-for-age shows more sensitivity to short-term interventions compared to height-based indicators[25]. However, some studies have demonstrated significant impacts on weight-for-age indicators. The Zimbabwe Sanitation Hygiene Infant Nutrition Efficacy (SHINE) trial found that WASH interventions combined with improved infant and young child feeding resulted in significant improvements in weight-for-age z-scores, particularly among children aged 12-24 months [14]. Similarly, the WASH Benefits trials demonstrated some positive trends in weight-based indicators when interventions were implemented with high fidelity[26].

The trend toward improvement in the intervention group, while not statistically significant, may suggest that SLTS interventions can influence weight-for-age through reduced morbidity from diarrheal diseases and other infections. The lack of statistical significance may be related to reasons such as the relatively short period of the intervention and the lack of a nutrition specific complementary intervention.

While valuable for improving sanitation behaviors, the SLTS intervention had no impact on the nutritional outcomes (wasting and underweight) of children aged 6-59 months. These findings suggest that for SLTS to have a meaningful impact on wasting and underweight, it should be integrated with nutrition-specific interventions, such as improved infant and young child feeding practices, micronutrient supplementation, and enhanced healthcare services. Long-term follow-up studies, spanning one to two years post-intervention, are also needed to assess the sustained impact of SLTS on nutritional outcomes in children aged 6-59 months.

There was a significant reduction in diarrheal disease prevalence in the intervention group, decreasing from 12.7% to 5.9%, compared to the control group, where prevalence remained relatively similar (13.6% to 13.2%). This difference was statistically significant at endline (*X*^2^=6.738, df=1, p=0.009), representing a 53% reduction in diarrheal episodes. These findings are consistent with other previous studies demonstrating the effectiveness of school-based sanitation interventions in reducing diarrheal diseases. A quasi-experimental study in South-Western Ethiopia found that Community-Led Total Sanitation and Hygiene (CLTSH) implementation significantly reduced diarrheal diseases in children under five years of age [27] Similarly, a propensity scores matched analysis across multiple low and middle-income countries demonstrated significant impacts of improved sanitation on diarrhea reduction among rural under-five children [10]. Additionally, a cluster randomized trial in Kenya that assessed the effect of school-based WASH programs reported significant reductions in diarrheal episodes among school-aged children and their younger siblings[13].

However, some studies have reported different findings. A large cluster-randomized trial in rural Mali found that villages receiving a behavioral, subsidy-free community-led sanitation intervention experienced diarrhea prevalence similar to control villages, despite substantial gains in latrine access and reductions in open defecation [28]. Similar null effects on diarrhea were also observed in other trials, including the rural Odisha (India) Sanitation Programme [29] and the WASH Benefits trials in Kenya and Bangladesh [26,30].

The significant reduction in diarrheal disease can be attributed to multiple components of the SLTS intervention. The comprehensive approach, including capacity building, formation of school health clubs, triggering activities, and continuous household monitoring, likely created multiple pathways for behavior change. These components led to increased awareness and behavior change in hygiene practices, such as handwashing and safe faeces disposal, which are critical in breaking the transmission routes of diarrheal pathogens. In addition, school children, trained as agents of change, effectively transmitted hygiene messages to households, further augmenting the effect of the intervention. Notably, the continuous monitoring through home visits by the study team, the area Public Health Officer, and Community Health Promoters may have also reinforced desirable sanitation and hygiene behaviors that support diarrhea incidence reduction.

### 4.1 Conclusion

The study found no significant improvement in nutritional outcomes (wasting and underweight) associated with the School-Led Total Sanitation (SLTS) intervention. This finding underscores the complex, multi-factorial nature of chronic malnutrition, which likely requires broader, long-term nutritional and health interventions beyond sanitation and hygiene alone. However, the intervention was associated with a significant 54% reduction in diarrhea prevalence in the intervention arm. This highlights the effectiveness of the School-Led Total Sanitation approach in addressing the problem of diarrhea, particularly in rural settings.

### 4.2 Recommendations

Given the demonstrated effectiveness in reducing diarrheal diseases, policymakers should consider integrating SLTS approaches into national sanitation and hygiene strategies. The Ministry of Health and Ministry of Education should collaborate to develop standardized SLTS implementation guidelines that can be scaled across different counties and contexts. This integration should include budget allocations, training protocols, and monitoring frameworks that ensure sustainable implementation.

SLTS programs should be designed as part of comprehensive packages that include nutrition-specific interventions such as infant and young child feeding counseling, micronutrient supplementation, and growth monitoring. This integrated approach is more likely to achieve meaningful impacts on nutritional outcomes while maintaining the proven effectiveness in diarrheal disease reduction.

### 4.3 Further Research

Future research should prioritize longitudinal studies with extended follow-up periods (2-5 years) to better understand the sustainability of behavior change and health improvements achieved through SLTS interventions. These studies should investigate whether the observed reductions in diarrheal diseases are maintained over time and whether longer exposure periods result in measurable nutritional improvements.

In this study, SLTS was implemented as a package of several elements. The impact of each individual element on the key outcomes was not evaluated. Future studies should therefore unbundle the intervention package and evaluate both the individual and combined effects of its elements. Generating such evidence will be critical for identifying the most cost-effective components, enabling policymakers to prioritize them in future implementation phases and optimize resource allocation.

## Data Availability

The authors will make available any requested data as will be required for the publication of this article

## Acknowledgement

We are profoundly grateful to God for His protection and mercies throughout this research. Our deepest appreciation goes to the Baringo County Health and Education Departments for providing all the necessary support, and to Mrs. Janet Chirchir, the Kiptoim area Public Health Officer, for her immense support. We extend our sincere thanks to the Headteachers of all participating schools, our dedicated Research Assistants, and all the respondents. Finally, we acknowledge our families for providing the nurturing and conducive environment that made this work possible.

## Declaration of generative AI and AI-assisted technologies in the writing process

During the preparation of this work, the author(s) used Grammarly to improve the grammar and sentence structure of the manuscript. After using this tool, the authors reviewed and edited the content as needed and take full responsibility for the content of the publication.

